# The impact of music on health-related quality of life, as quantified by the SF-36: A systematic review and meta-analysis

**DOI:** 10.1101/2021.11.30.21267066

**Authors:** J. Matt McCrary, Eckart Altenmüller, Clara Kretschmer, Daniel S. Scholz

**Affiliations:** Institute of Music Physiology and Musicians’ Medicine, Hannover University of Music, Drama and Media, Hannover, Germany; Prince of Wales Clinical School, University of New South Wales, Sydney, Australia

## Abstract

**Background/Objectives:** Increasing evidence supports the ability of music to broadly promote wellbeing and health-related quality of life (HRQOL). However, the magnitude of music’s effects on HRQOL is still unclear, particularly relative to established interventions, limiting inclusion of music interventions in health policy and care. The SF-36 is the most widely used instrument to evaluate HRQOL, with broad validity in evaluating the effects of a range of interventions. This study aims to synthesize and contextualize the impact of music interventions on HRQOL, as assessed by the SF-36.

**Methods:** MEDLINE; EMBASE; Web of Science; PsycINFO, clinicaltrials.gov, and ICTRP databases were searched on 30 July 2021. Randomized and single-group studies of music interventions which reported SF-36 data at pre- and post-intervention timepoints were included. Observational studies were excluded. The quality and certainty of evidence provided by included articles and meta-analysis results was appraised using GRADE. Inverse variance random effects meta-analyses quantified changes in SF-36 mental and physical component summary scores (respectively, ‘MCS’ and ‘PCS’) pre- to post-intervention and vs. common control groups.

**Results:** Analyses included 764 participants from 25 studies. Music interventions (music listening – 10 studies; music therapy – 7 studies; singing – 7 studies; gospel music – 1 study) significantly improved MCS (Mean difference (MD) [95% confidence interval]=3.0 [1.4, 4.6]; p<.001) and PCS (MD=1.0 [0.1, 2.0; p<.04) scores. In a subgroup (8 studies; music group – N=254; control – N=257) addition of music to standard treatment for a range of conditions significantly improved MCS scores vs. standard treatment alone (MD=3.7 [0.4, 7.1; p<.03). Effects did not vary between music listening, therapy and singing intervention types or doses (*p≥*.*12*); no evidence of small study or publication biases was present in any analysis (*p≥*.*31*). Music’s impact on MCS scores meets SF-36 minimum important difference thresholds (MD≥3) and is within the range of established interventions.

**Conclusions:** This study provides Moderate quality evidence that music interventions can generally be used to provide clinically meaningful improvements in HRQOL. Further study is needed to determine optimal music interventions and doses for distinct clinical and public health scenarios.

**Funding:** Alexander von Humboldt Foundation

**Registration:** PROSPERO (ID: CRD42021276204)

## Introduction

Health-related quality of life (HRQOL) is a broad concept capturing ‘an individual’s or group’s perceived physical and mental health over time.’^1^ HRQOL is closely related to and frequently used interchangeably with ‘well-being’,^1^ with the importance of these broad health concepts reflected by their prominence in United Nations Sustainable Development Goal #3: *‘To ensure healthy lives and promote well-being for all at all ages’*.^2^

Listening to and making music (*e.g. by singing or playing instruments*) is increasingly advocated, including in a recent World Health Organization report, as a means of improving HRQOL and various domains of well-being in clinical and healthy populations.^3-7^ However, a lack of clarity regarding the magnitude of music effects on HRQOL, particularly compared to other established health interventions, presents clear challenges to the inclusion of music in health policies and care at local, national and international levels.^8,9^ Additionally, optimal music intervention types and doses for specific scenarios are still unclear.^3,9^ In short, evidence supporting music’s impact on HRQOL is presently broad and narrative, while specific, quantitative evidence is needed to justify an increased use of music interventions in health policy and care.

The Short Form (SF)-36 HRQOL questionnaire is the most widely used patient-reported outcome instrument in health research, demonstrating strong validity, sensitivity, and reliability across a range of languages, interventions and clinical and healthy populations.^10-12^ The SF-36 has also been frequently used in studies of music interventions,^3,4^ providing a means of both quantifying and easily contextualising the magnitude of music’s effects on HRQOL using a broadly valid and applicable instrument. Accordingly, the aim of this study is to quantitatively synthesize and contextualize the impact of music interventions on HRQOL using the SF-36. A secondary study aim is to evaluate the relative impact of music intervention types and doses on HRQOL.

## Methods

### Review registration

The protocol for this systematic review and meta-analysis was prospectively registered with PROSPERO (ID: CRD42021276204).

### Search Strategy

Four databases – MEDLINE; EMBASE; Web of Science; PsycINFO – and three clinical trials registries – Cochrane Central Register of Controlled Trials (CENTRAL); clinicaltrials.gov; International Clinical Trials Registry Platform (ICTRP) – were searched for peer-reviewed articles, clinical trial registrations and ‘grey’ research reports on 30 July 2021 using the following query:

> (Music* OR singing OR listening) AND (SF12 OR SF36 OR SF-12 OR SF-36 OR ‘short form 36’ OR ‘short form 12’)

All related subject headings were included where possible and no limitations on study date or language were imposed. The reference lists of included studies and relevant systematic reviews were also hand searched for additional relevant studies.

### Article screening and inclusion/exclusion criteria

Following removal of duplicate records, titles and abstracts of database search results were screened, followed by full-text review of potentially relevant abstracts against inclusion/ exclusion criteria. Screening and full-text review were performed independently in duplicate by two study authors (JMM and CK), with disagreements resolved through discussion.

Inclusion criteria were randomized and non-randomized studies investigating the impact of music-making (*e.g. instrumental music; singing; ‘active’ music therapy*) and/or music listening (*e.g. to recorded or live music; ‘receptive’ music therapy*) interventions on HRQOL in adults using the SF-36 or SF-12 (reduced version) questionnaires. No restrictions were made on eligible control groups. Studies that investigated the impact of music on HRQOL as either a primary or secondary objective were both eligible for inclusion. Additionally, studies must have reported the SF-36/SF-12 Mental Component Summary (MCS) score and/or Physical Component Summary (PCS) score, or data enabling the calculation of a MCS and/or PCS score (*e.g. data from all 8 SF-36/SF-12 subscales*), at both pre-intervention and immediate post-intervention timepoints. Higher MCS and PCS scores indicated better mental and physical HRQOL, respectively. MCS and PCS from the SF-12 and SF-36 have demonstrated good consistency across a range of populations.^13,14^

MCS and PCS scores are both calculated using norm-based scoring methods including all 8 subscales of the SF-36/12: Physical Functioning; Role Physical; Bodily Pain; General Health; Vitality; Social Functioning; Role-Emotional; Mental Health. MCS scores are more heavily weighted towards Vitality, Social Functioning, Role-Emotional, and Mental Health subscale scores. PCS scores are more heavily weighted towards Physical Functioning, Role-Physical, Bodily Pain and General Health subscale scores.^15^

Exclusion criteria were observational and cross-sectional studies and studies which investigated other music-related activities that do not focus on music-making or listening (*e.g. songwriting*).

### Study and Evidence Appraisal (GRADE)

The quality of evidence supporting review conclusions was appraised using the GRADE system. GRADE provides a framework for evaluating the risk of bias of individual studies, as well as the level of certainty supporting specific review results.^16^ GRADE was selected for this review because of its broad applicability to different study types and because it has been “designed for reviews…that examine alternative management strategies.”^16^

The risk of bias of individual studies was evaluated using the following standard criteria: allocation concealment; blinding (of assessors & data analysts); % lost to follow-up; intention to treat analysis; selective outcome reporting; use of individual randomization; control for carryover effects (crossover study design).^17^ Based on these criteria, an evidence quality rating of High, Moderate, Low, or Very Low was assigned to each study using established procedures.^17^ All studies were appraised independently by two study authors (JMM and CK), with any disagreements resolved through discussion. The overall quality and certainty of evidence supporting the results of each meta-analysis was then appraised by the primary author in consultation with the authorship team using the same rating scale.^17^

### Data extraction

Demographic, music and control intervention, and pre- and post-intervention SF-36/SF-12 data were extracted in duplicate by two study authors (JMM and CK). Data from all available SF-36/SF-12 subscales were extracted, as well as MCS and PCS summary scores. To maximize consistency of data across studies, MCS and PCS scores were recalculated where possible from underlying subscale data using the methodology of Ware, Kosinsky and Keller.^15^ Missing MCS and PCS standard deviations were imputed from Mental Health and Physical Function subscale scores, respectively, or median/minimum/maximum/interquartile range data as per established methods.^18,19^ Authors of studies meeting inclusion criteria but reporting unclear or incomplete SF-36/SF-12 data were contacted to retrieve compatible MCS and/or PCS data.

### Data analysis and statistics (including meta-analysis)

Weighted inverse variance random effects meta-analyses were conducted to determine the aggregate pre- to post-intervention change in MCS and PCS scores. Additionally, inverse variance random effects meta-analyses were performed on post-intervention MCS and PCS scores in music vs. control groups common to at least 3 studies. The presence of statistical heterogeneity, indicating significant variation in the overall effects of music interventions on MCS and PCS scores, was evaluated using the χ^2^ test and I^2^ statistic. Potential small study/ publication biases were evaluated using Egger’s test.^20^ Sensitivity analyses were performed where possible according to music intervention types (*e.g. music therapy, singing, music listening*) and quality of study evidence (*very low/low vs. moderate/high*). Additionally, exploratory non-parametric Spearman correlation analyses were performed to evaluate potential links between key characteristics of the music intervention ‘dose’ (*intervention duration; music session frequency and length*) and MCS and PCS scores. Significance was set at α=0.05 for all statistical tests except meta-analysis main effects – α=0.033 was used for meta-analysis main effects to control for multiplicity of related MCS and PCS outcomes.^21^ Analyses were conducted in RevMan v5.4 (Cochrane Collaboration; London, United Kingdom) and SPSS v26 (IBM; Armonk, NY, USA).

Finally, published meta-analyses of MCS and PCS scores from established non-pharmaceutical/medical health interventions were retrieved to serve as a basis for comparison with results of the present study.

## Results

Data from 25 eligible studies and N=764 total participants (*mean age = 60* (*standard deviation=11 years*)) were included in the present study (Figure 1)(*see Supplementary Appendix for full details of excluded studies*). Included studies were conducted in Australia,^22^ Brazil,^23-25^ China (*Hong Kong SAR*),^26^ Germany,^27^ India,^28^ Italy,^29-31^ The Netherlands,^32,33^ Spain,^34^ Sweden,^35^ Thailand,^36^ Turkey,^37^ the United Kingdom,^38-41^ and the United States.^42-46^ Included studies were comprised of: 21 investigations of clinical populations and 4 of healthy populations; 10 investigations of music listening, 7 music therapy, 7 singing, and 1 ‘gospel music’ intervention; 19 RCTs and 6 single-group studies; 8 RCTs included comparisons to a ‘treatment as usual’ control group, 3 RCTs used meditation control groups, and 8 RCTs used a range of other disparate control groups. MCS and PCS scores were available for 24 studies; MCS data only was available in one additional study.^45^

**Figure 1.**
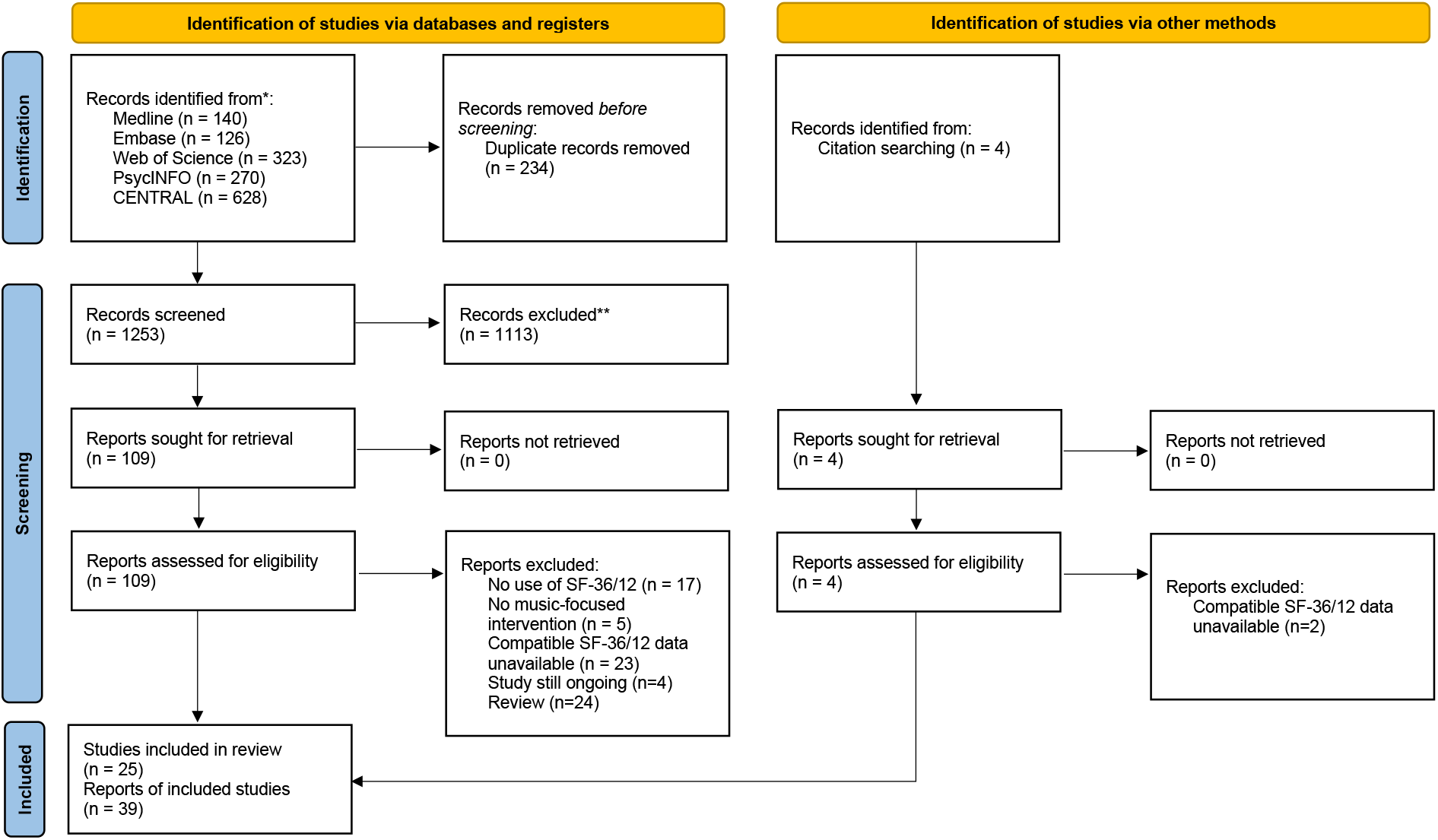
PRISMA flow diagram detailing the results of record screening and exclusion procedures.^61^

Evidence quality was High in 5 studies, Moderate in 10 studies, Low in 7 studies, and Very Low in 3 studies (*see Supplementary Appendix for further details regarding GRADE ratings for each study*).

### Pre-post changes

Music interventions significantly increased both MCS and PCS scores from pre-intervention values by an average of 3.0 (*p<.001*) and 1.0 (*p=.032*) points, respectively (*Standardized mean differences [95% confidence interval]: MCS=0.25[0.14,0.36] PCS=0.15[0.05,0.25])* (Figures 2 and 3). MCS scores (*N=764 participants*) were significantly greater in Moderate/ High vs. Very Low/Low quality studies (*p=.03*) and varied significantly across intervention types (*p=.03*)(Figure 2). However, MCS scores did not significantly vary across intervention types when the one ‘gospel music’ intervention study^42^ was excluded (*p=0.12*). PCS scores (*N=748 participants*) did not significantly vary according to study quality (*p=0.30*) or intervention type (*p=0.26*).

**Figure 2.**
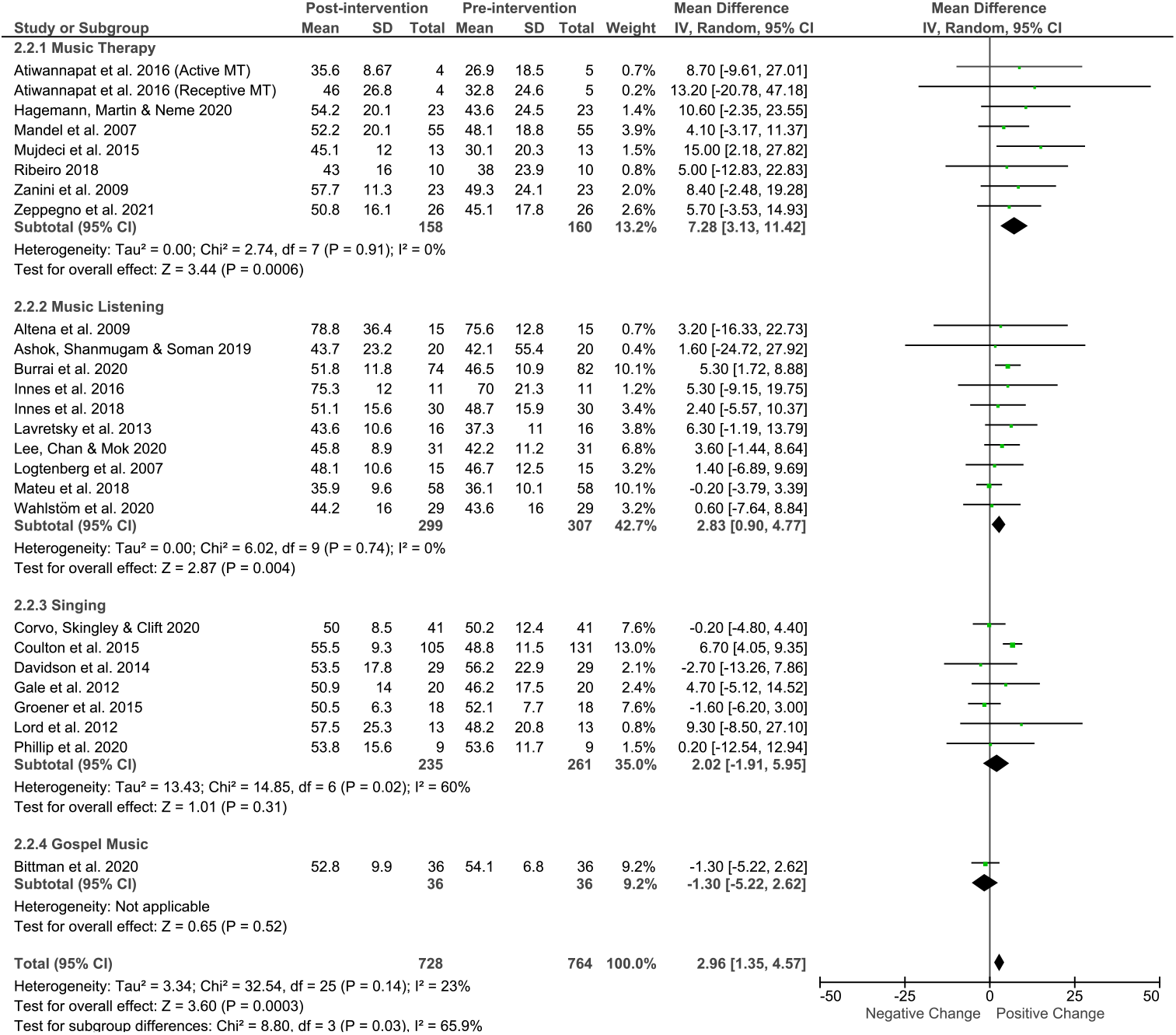
Meta-analysis of pre- to post-intervention effects of music interventions on SF-36 MCS scores, stratified by music intervention type. IV = ‘inverse variance’. ‘Total’ refers to the total number of participants included in analyses at pre- and post-intervention timepoints.

**Figure 3.**
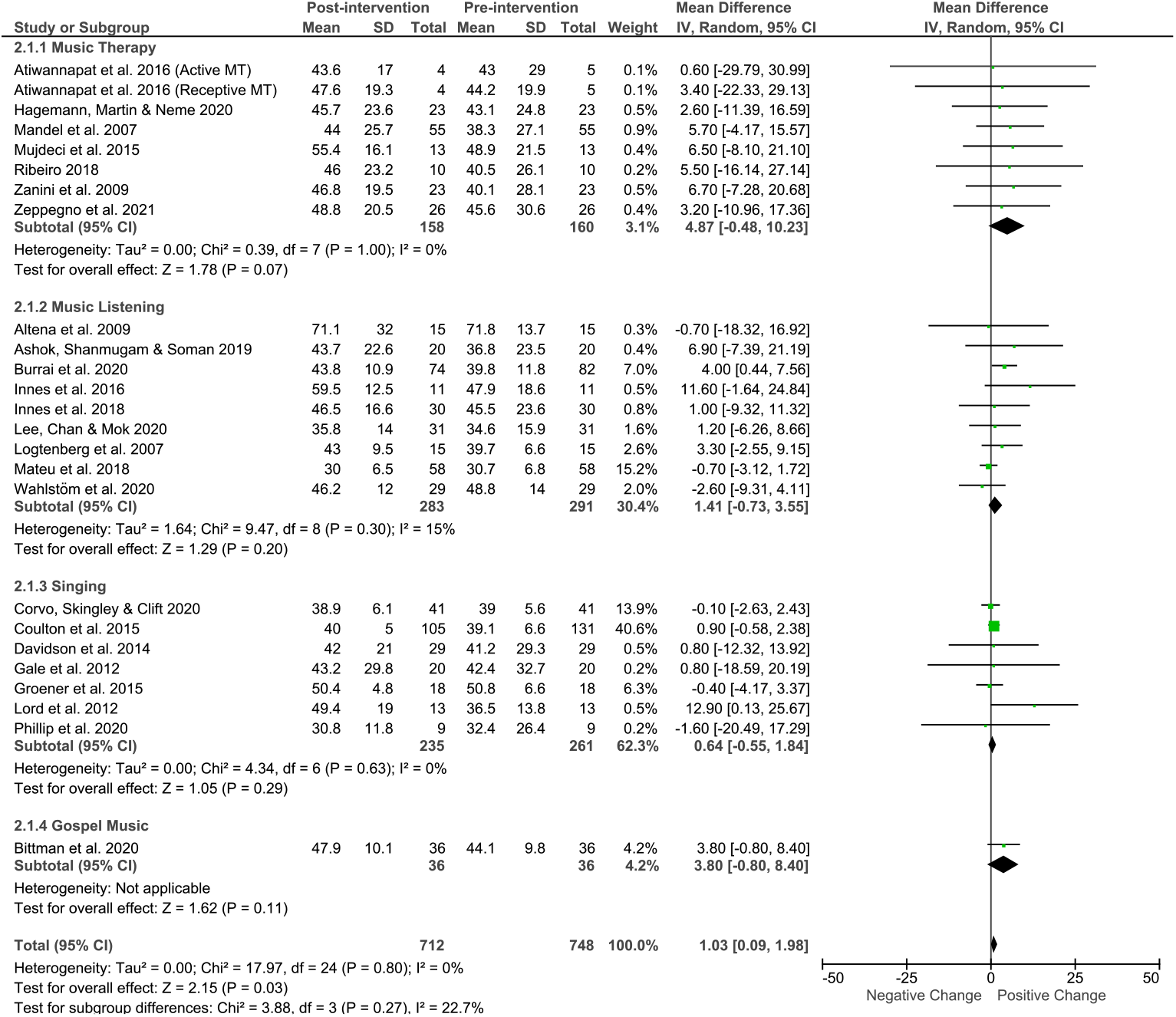
Meta-analysis of pre- to post-intervention effects of music interventions on SF-36 PCS scores, stratified by music intervention type. IV = ‘inverse variance’. ‘Total’ refers to the total number of participants included in analyses at pre- and post-intervention timepoints.

No key characteristics of the music intervention ‘dose’ (*intervention duration; music session frequency and length*) were significantly correlated with MCS or PCS scores (*p>.32*). No significant statistical heterogeneity (*MCS: p=0.12; PCS: p=0.80*) or evidence of small study/publication bias (*MCS: p=0.31; PCS: p=0.70*)(*see Supplementary Appendix for further details*) was present in either analysis. Results of these meta-analyses were also judged to be minimally affected by individual study biases but limited by the imprecision of relatively wide confidence intervals. Accordingly, results are appraised to provide Moderate quality evidence, indicating that ‘*the true effect is probably close to the estimated effect*’.^16^

### Music + Treatment as Usual vs. Treatment as Usual alone

Adding music interventions to Treatment as Usual (TAU) significantly increased MCS scores vs. TAU alone by an average of 3.7 points (*p=.029; standardized mean difference [95% confidence interval] = 0.24[0.02,0.45])(Music+TAU – N=254 participants; TAU alone – N=257)*(Figure 4). Similar effects were not observed for PCS scores (*p=.16*)(*Music+TAU – N=254 participants; TAU alone – N=257*)(Figure 5). Improvements in MCS in music + TAU vs. TAU groups did not vary significantly with study quality or music intervention type (*p>.28*). No significant statistical heterogeneity (*MCS: p=.26; PCS: p=.78*) or evidence of small study/publication biases (*MCS: p=.72; PCS: p=.64*) (*see Supplementary Appendix*) was present in either analysis. Results of these meta-analyses were judged to be minimally affected by individual study biases but limited by the imprecision of wide confidence intervals. Accordingly, results are appraised to provide Moderate quality evidence.

**Figure 4.**
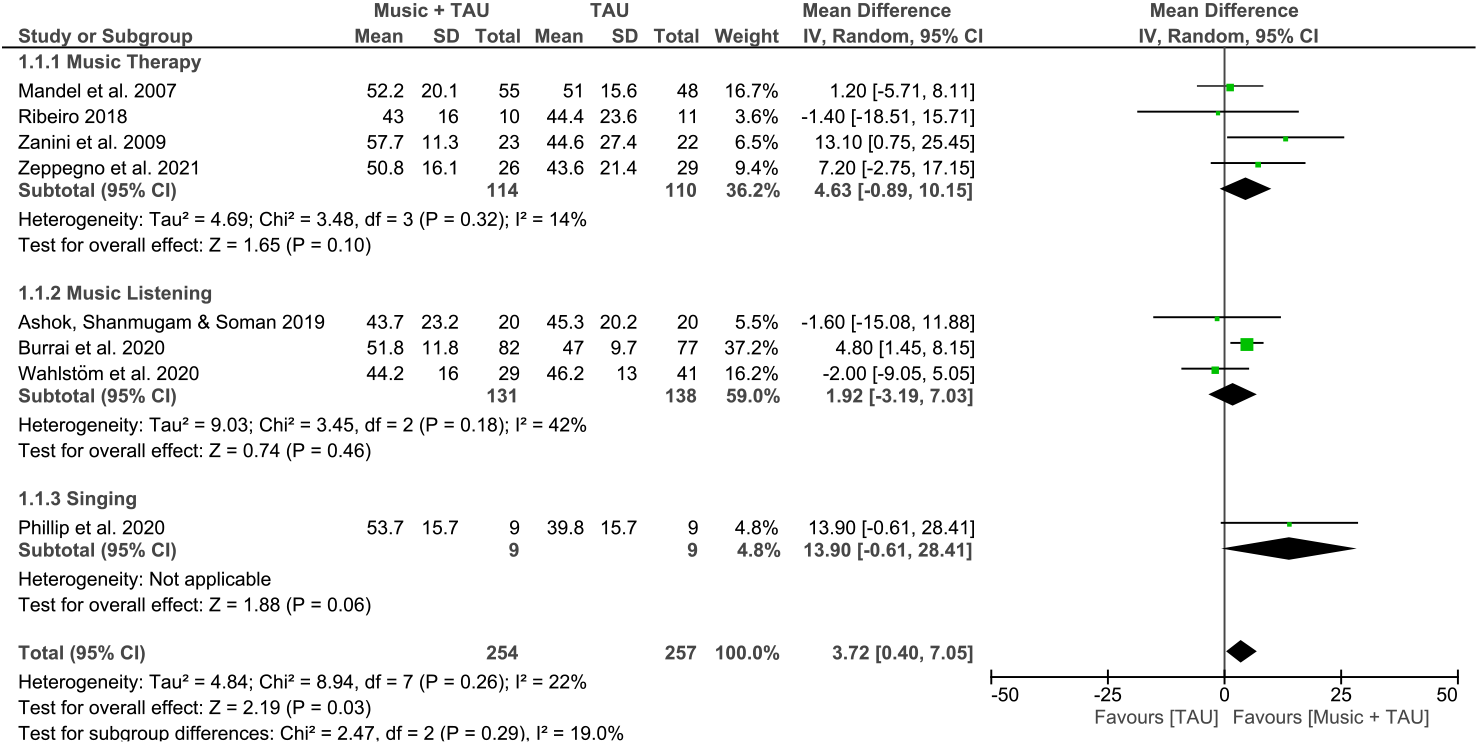
Effects of music interventions added to Treatment as Usual (TAU) vs. TAU alone on SF-36 MCS scores, stratified by music intervention type. IV = ‘inverse variance’. ‘Total’ refers to the total number of participants included in analyses at pre- and post-intervention timepoints.

**Figure 5.**
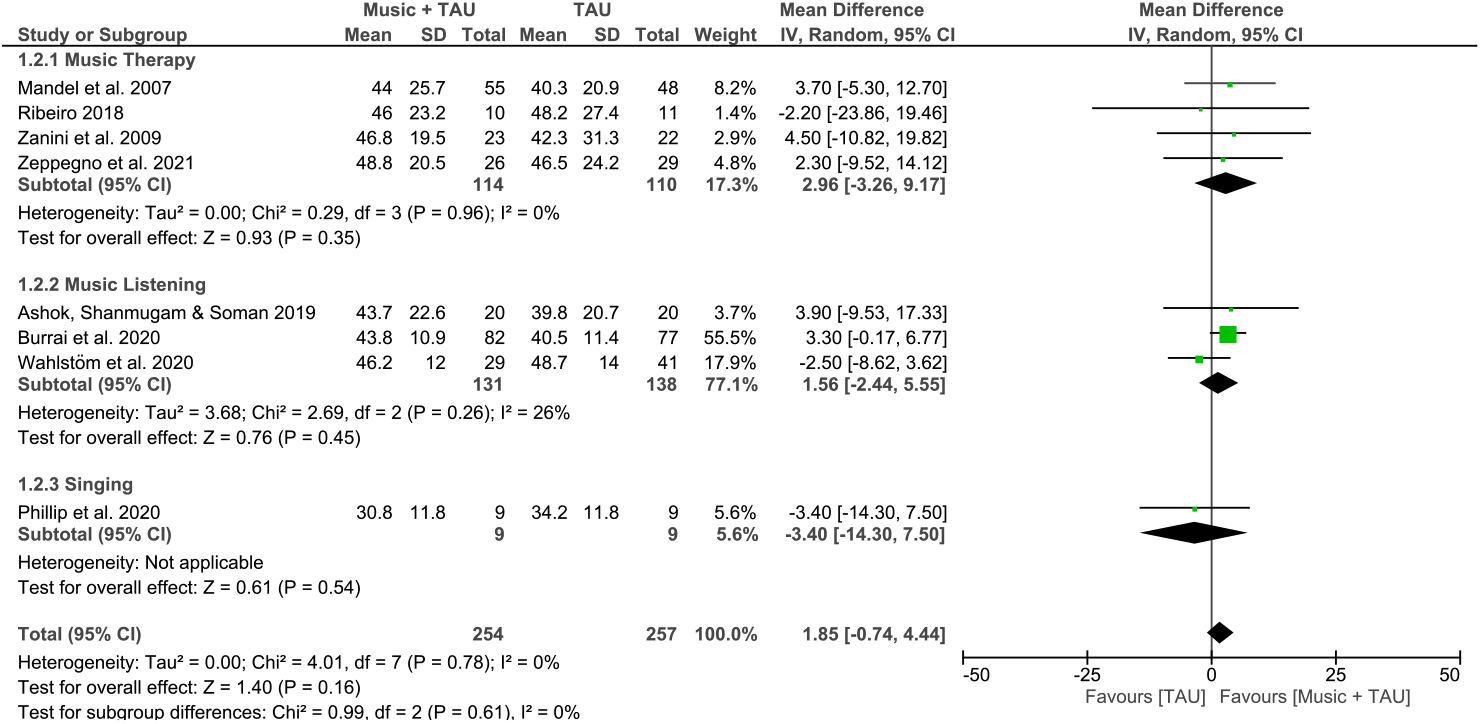
Effects of music interventions added to Treatment as Usual (TAU) vs. TAU alone on SF-36 PCS scores, stratified by music intervention type. IV = ‘inverse variance’. ‘Total’ refers to the total number of participants included in analyses at pre- and post-intervention timepoints.

### Music listening vs. meditation

No significant differences in MCS or PCS scores in music listening vs. meditation intervention studies were present across three included studies (*p>.30*)(*Music listening – N=57; Meditation – N=64)*(*see Supplementary Appendix*). Once again, no significant statistical heterogeneity or evidence of small study/publication biases was present in either analysis (*p>.34*). Results are, however, limited by the small number of studies and wide confidence intervals and judged to provide Low quality evidence – ‘*the true effect might be markedly different from the estimated effect’*.^16^

### Music effects in context

Effects of music interventions pre-post and vs. TAU on MCS scores meet or exceed, respectively, the proposed 3-point minimum important difference threshold for MCS and PCS scores.^15^ Pre-post changes in PCS scores (*1 point improvement*) fall below this threshold. Music intervention effects on MCS scores are similar to the impact of weight loss on PCS scores in studies of obese adults (*2.8 point improvement; no significant MCS change*).^47^ However, the magnitude of music intervention effects on both MCS and PCS scores is substantially smaller than the effects of resistance exercise (*i.e. strength training*) in older adults from mixed clinical and healthy populations vs. mixed control groups (*standardized mean difference – MCS=0.54, PCS=0.50)*^*48*^ and mixed modes of exercise in participants with knee osteoarthritis vs. inactive or psycho-educational control groups (*standardized mean difference – MCS=0.44, PCS=0.52*).^49^

## Discussion

This meta-analysis of 25 studies of music interventions provides clear and quantitative Moderate quality evidence that music interventions have a clinically significant impact on mental HRQOL. Additionally, a subset of 8 studies demonstrated a clinically significant benefit to mental HRQOL of adding music interventions to usual treatment for a range of conditions. The impact of music interventions on physical HRQOL is substantially smaller and of potentially equivocal practical importance.^15^

Included studies presented considerable heterogeneity in study populations and geographic locations, music intervention types and doses, and TAU control groups. However, no statistical heterogeneity or evidence of small study or publication bias was present in any analyses. This suggests that results approximate the true, albeit general, effects of music interventions on HRQOL. Further research is still needed to provide guidance regarding optimal music interventions and doses in distinct clinical and public health scenarios.

The effects of music interventions on MCS scores (*pre-post and music+TAU vs. TAU*) are within the range, albeit on the low end, of established non-pharmaceutical/medical,^47-50^ as well as medical/pharmaceutical,^51-53^ health interventions and likely to be clinically significant.^15^ Accordingly, this review quantitatively confirms previous narrative literature syntheses asserting that music interventions can produce meaningful improvements in wellbeing and HRQOL.^3-6^ Of particular interest for future study and health policy is the fact that these benefits are achieved through a broadly rewarding activity.^54^ While uptake and adherence challenges persist with other non-pharmaceutical/medical interventions (*e.g. weight loss, exercise*),^55,56^ music is ‘reliably ranked as one of life’s greatest pleasures.’^57^ As such, music interventions may present a more attractive and effective non-pharmaceutical alternative to other health interventions. Further study is required to investigate this hypothesis and clarify the specific utility of music vs. other established interventions.

Additional, targeted research is also needed to provide insights into the mechanisms of music interventions’ positive impacts on HRQOL – the *who/what/when/where/how* underpinning their effectiveness. The absence of any significant differences between music intervention types and doses in the present analyses is intriguing but not definitive; these results could also be simply explained by the diversity of included populations and interventions, even within specific intervention types (*particularly clearly demonstrated for ‘music listening’ interventions in Table 1*). Broad confidence intervals of both main and intervention type-specific effects likely also reflect the influence of diverse interventions. A recent analysis indicates that the mechanisms of music’s impact on health are complex and specific to distinct settings, suggesting that targeted study is required to determine optimal music intervention characteristics in each setting.^58^ However, other analyses propose that such targeted research may be able to be rapidly generalized to other settings if foundational physiologic mechanisms of music intervention effects can be identified and targeted.^59^

**Table 1.**
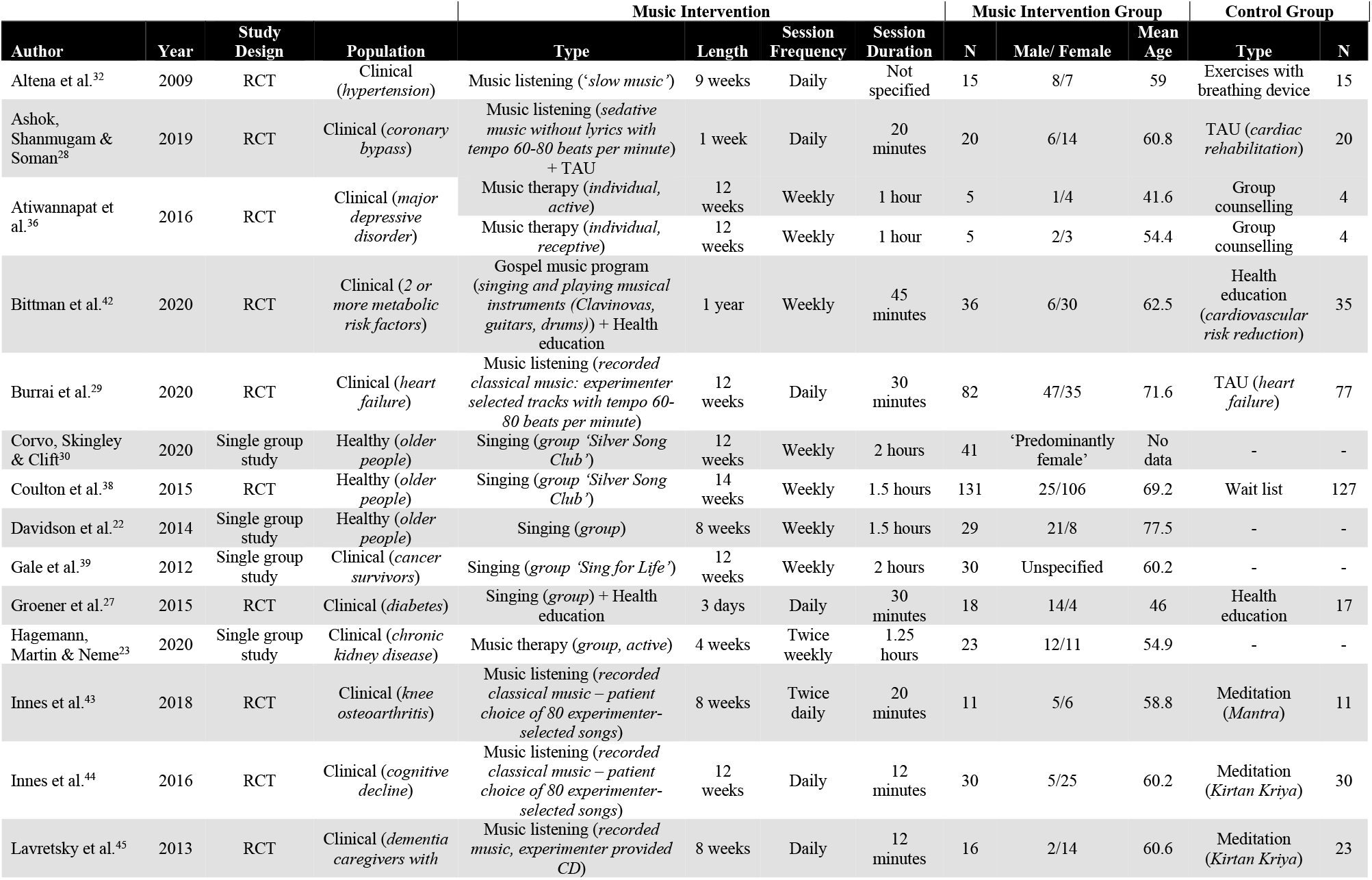
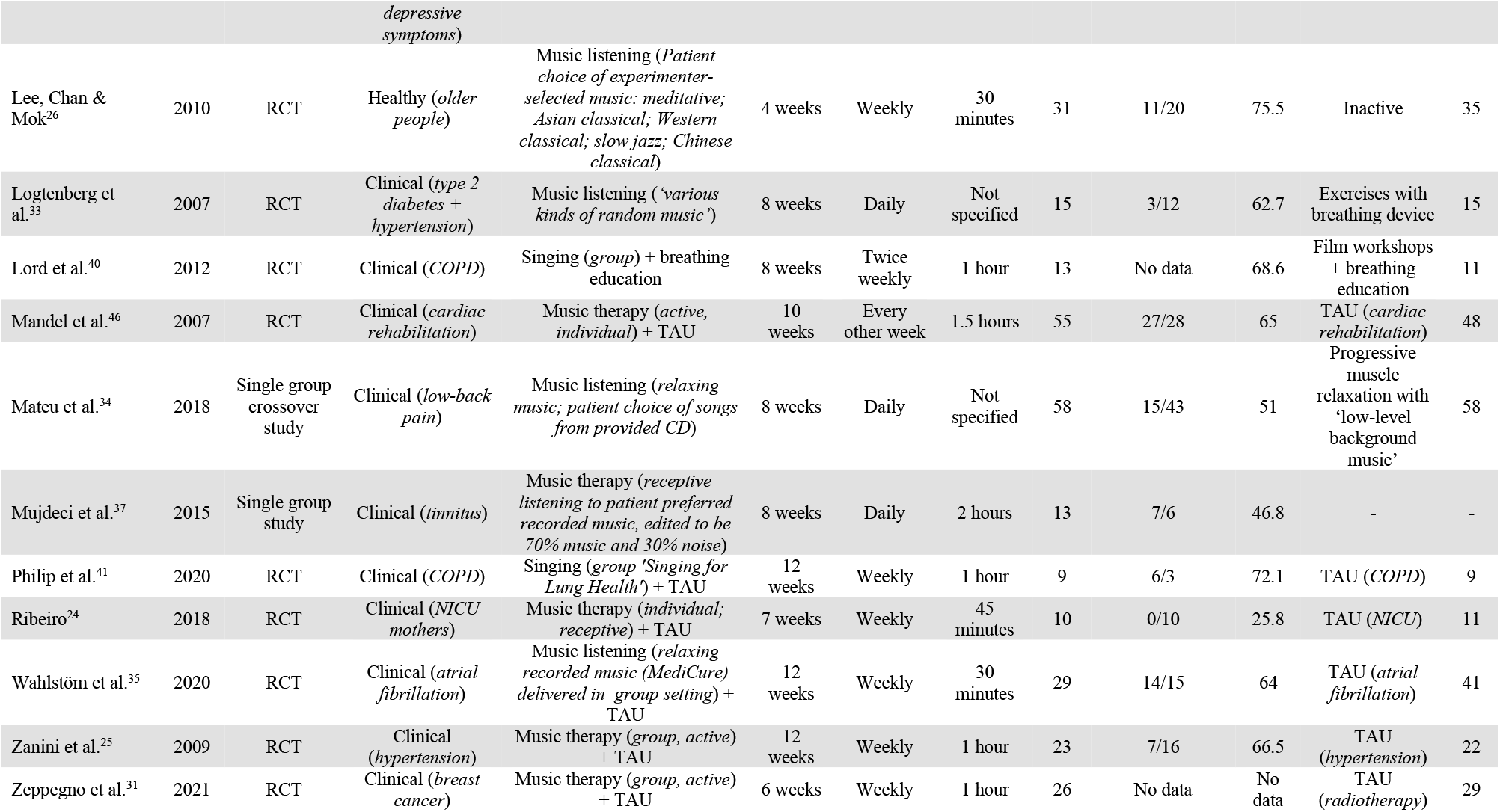
Characteristics of included studies. RCT= randomized controlled trial; TAU = treatment as usual; COPD = chronic obstructive pulmonary disorder; NICU = neonatal intensive care unit.

This review is limited by its broad inclusion criteria which limits conclusions regarding the effects of specific music interventions in particular scenarios, especially given the diversity of included interventions. Additionally, standardized mean differences describing the magnitude of pre-post intervention effects have been shown to be prone to bias and must be interpreted with caution.^60^ However, the similar effect size of music interventions on mental HRQOL in Music+TAU vs. TAU analyses provides additional confidence in the magnitude of pre-post MCS changes. Finally, this review is ultimately limited to studies evaluating the impact of music interventions HRQOL using the SF-36 or SF-12 instruments, a possibly skewed subset of music intervention studies. Statistical homogeneity, the absence of apparent publication/ small study biases, and the broad psychometric rigor of the SF-36 and SF-12^13,14^ suggest that results of the present review approximate the true effect of music interventions on HRQOL. However, the possibility remains that this subset of studies is not representative of music’s general effects on HRQOL or that the SF-36/12 instruments do not completely capture the impact of music on HRQOL. This uncertainty is reflected in the Moderate quality rating of key review results, indicating that ‘the true effect is probably close to the estimated effect’.^16^

## Conclusions

This study provides Moderate quality quantitative evidence of a clinically significant impact of music interventions on mental HRQOL. The impact of music on physical HRQOL is potentially equivocal. The magnitude of music’s impact on mental HRQOL is within the range, albeit at the low end, of effects of established non-pharmaceutical/medical interventions (*e.g. exercise, weight loss*). Future research is needed to clarify optimal music interventions and doses for use specific clinical and public health scenarios.

## Supporting information

PRISMA

## Data Availability

All data produced in the present work are contained in the manuscript

## Acknowledgements

J. Matt McCrary is supported by a Postdoctoral Fellowship from the Alexander von Humboldt Foundation. The authors have no competing interests to declare.

## Data Availability

All review data is contained in the manuscript and supplementary appendix.

## References

1. Centers for Disease Control and Prevention. Health-related quality of life (HRQOL). Centers for Disease Control and Prevention. (https://www.cdc.gov/hrqol/index.htm).

2. United Nations. 2030 Agenda for Sustainable Development. New York: United Nations, 2015.

3. McCrary JM, Redding E, Altenmüller E. Performing arts as a health resource? An umbrella review of the health impacts of music and dance participation. PloS one 2021;16(6):e0252956.

4. Daykin N, Mansfield L, Meads C, et al. What works for wellbeing? A systematic review of wellbeing outcomes for music and singing in adults. Perspectives in Public Health 2018;138(1):39–46.

5. World Health Organization. What is the evidence on the role of the arts in improving health and well-being? A scoping review: World Health Organization. Regional Office for Europe, 2019.

6. Fancourt D, Warran K, Aughterson H. Evidence Summary for Policy: The role of the arts in improving health & wellbeing. 2020.

7. Gordon-Nesbitt R, Howarth A. The arts and the social determinants of health: findings from an inquiry conducted by the United Kingdom All-Party Parliamentary Group on Arts, Health and Wellbeing. Arts & health 2020;12(1):1–22.

8. Clift S, Phillips K, Pritchard S. The need for robust critique of research on social and health impacts of the arts. Cultural Trends 2021:1–18. DOI: 10.1080/09548963.2021.1910492.

9. Bickerdike L, Booth A, Wilson PM, Farley K, Wright K. Social prescribing: less rhetoric and more reality. A systematic review of the evidence. BMJ open 2017;7(4).

10. Scoggins JF, Patrick DL. The use of patient-reported outcomes instruments in registered clinical trials: evidence from http://ClinicalTrials.gov. Contemporary clinical trials 2009;30(4):289-292.

11. McHorney CA, Ware Jr JE, Raczek AE. The MOS 36-Item Short-Form Health Survey (SF-36): II. Psychometric and clinical tests of validity in measuring physical and mental health constructs. Medical care 1993:247–263.

12. Jenkinson C, Wright L, Coulter A. Criterion validity and reliability of the SF-36 in a population sample. Quality of Life Research 1994;3(1):7–12.

13. Luo X, George ML, Kakouras I, et al. Reliability, validity, and responsiveness of the short form 12-item survey (SF-12) in patients with back pain. Spine 2003;28(15):1739–1745.

14. Jenkinson C, Chandola T, Coulter A, Bruster S. An assessment of the construct validity of the SF-12 summary scores across ethnic groups. Journal of Public Health 2001;23(3):187–194.

15. Ware J, Kosinski M, Keller S. SF-36 physical and mental health summary scales. A user’s manual 2001:1994.

16. Guyatt G, Oxman AD, Akl EA, et al. GRADE guidelines: 1. Introduction—GRADE evidence profiles and summary of findings tables. Journal of clinical epidemiology 2011;64(4):383–394.

17. Balshem H, Helfand M, Schünemann HJ, et al. GRADE guidelines: 3. Rating the quality of evidence. Journal of clinical epidemiology 2011;64(4):401–406.

18. Matcham F, Scott IC, Rayner L, et al. The impact of rheumatoid arthritis on quality-of-life assessed using the SF-36: a systematic review and meta-analysis. Seminars in arthritis and rheumatism: Elsevier; 2014:123–130.

19. Wan X, Wang W, Liu J, Tong T. Estimating the sample mean and standard deviation from the sample size, median, range and/or interquartile range. BMC medical research methodology 2014;14(1):1–13.

20. Egger M, Smith GD, Schneider M, Minder C. Bias in meta-analysis detected by a simple, graphical test. Bmj 1997;315(7109):629–634.

21. Jakobsen JC, Wetterslev J, Winkel P, Lange T, Gluud C. Thresholds for statistical and clinical significance in systematic reviews with meta-analytic methods. BMC medical research methodology 2014;14(1):1–13.

22. Davidson JW, McNamara B, Rosenwax L, Lange A, Jenkins S, Lewin G. Evaluating the potential of group singing to enhance the well-being of older people. Australasian Journal on Ageing 2014;33(2):99–104.

23. Hagemann PdMS, Martin LC, Neme CMB. The effect of music therapy on hemodialysis patients’ quality of life and depression symptoms. Brazilian Journal of Nephrology 2018;41:74–82.

24. Ribeiro MKA. Influência da Intervenção Musicoterapêutica sobre a Variabilidade da Frequência Cardíaca e Aspectos Biopsicosociais em Mães de Prematuros: Estudo Randomizado. Ciências da Saúde: Universidade Federal de Goiás; 2019.

25. Zanini CRdO, Jardim PCBV, Salgado CM, et al. Music therapy effects on the quality of life and the blood pressure of hypertensive patients. Arquivos brasileiros de cardiologia 2009;93:534–540.

26. Lee YY, Chan MF, Mok E. Effectiveness of music intervention on the quality of life of older people. Journal of advanced nursing 2010;66(12):2677–2687.

27. Groener J, Neus I, Kopf S, et al. Group singing as a therapy during diabetes training–A randomized controlled pilot study. Experimental and Clinical Endocrinology & Diabetes 2015;123(10):617–621.

28. Ashok A, Shanmugam S, Soman A. Effect of Music Therapy on Hospital Induced Anxiety and Health Related Quality of Life in Coronary Artery Bypass Graft Patients: A Randomised Controlled Trial. Journal of Clinical & Diagnostic Research 2019;13(11).

29. Burrai F, Sanna GD, Moccia E, et al. Beneficial effects of listening to classical music in patients with heart failure: a randomized controlled trial. Journal of cardiac failure 2020;26(7):541–549.

30. Corvo E, Skingley A, Clift S. Community singing, wellbeing and older people: implementing and evaluating an English singing for health intervention in Rome. Perspectives in Public Health 2020;140(5):263–269.

31. Zeppegno P, Krengli M, Ferrante D, et al. Psychotherapy with Music Intervention Improves Anxiety, Depression and the Redox Status in Breast Cancer Patients Undergoing Radiotherapy: A Randomized Controlled Clinical Trial. Cancers 2021;13(8):1752.

32. Altena MR, Kleefstra N, Logtenberg SJ, Groenier KH, Houweling ST, Bilo HJ. Effect of device-guided breathing exercises on blood pressure in patients with hypertension: a randomized controlled trial. Blood pressure 2009;18(5):273–279.

33. Logtenberg SJ, Kleefstra N, Houweling ST, Groenier KH, Bilo HJ. Effect of device-guided breathing exercises on blood pressure in hypertensive patients with type 2 diabetes mellitus: a randomized controlled trial. Journal of hypertension 2007;25(1):241–246.

34. Mateu M, Alda O, Inda M-d-M, et al. Randomized, Controlled, Crossover Study of Self-administered Jacobson Relaxation in Chronic, Nonspecific, Low-back Pain. Alternative Therapies in Health & Medicine 2018;24(6).

35. Wahlström M, Rosenqvist M, Medin J, Walfridsson U, Rydell-Karlsson M. MediYoga as a part of a self-management programme among patients with paroxysmal atrial fibrillation–a randomised study. European Journal of Cardiovascular Nursing 2020;19(1):74–82.

36. Atiwannapat P, Thaipisuttikul P, Poopityastaporn P, Katekaew W. Active versus receptive group music therapy for major depressive disorder—A pilot study. Complementary therapies in medicine 2016;26:141–145.

37. Müjdeci B, Köseoglu S, Özcan İ, Dere H. Effect of music therapy on quality of life in individuals with tinnitus. MARMARA MEDICAL JOURNAL 2015;28(1):38–44.

38. Coulton S, Clift S, Skingley A, Rodriguez J. Effectiveness and cost-effectiveness of community singing on mental health-related quality of life of older people: randomised controlled trial. The British Journal of Psychiatry 2015;207(3):250–255.

39. Gale NS, Enright S, Reagon C, Lewis I, Van Deursen R. A pilot investigation of quality of life and lung function following choral singing in cancer survivors and their carers. Ecancermedicalscience 2012;6.

40. Lord VM, Hume VJ, Kelly JL, et al. Singing classes for chronic obstructive pulmonary disease: a randomized controlled trial. BMC Pulmonary Medicine 2012;12(1):1–7.

41. Philip KE, Lewis A, Jeffery E, et al. Moving singing for lung health online in response to COVID-19: experience from a randomised controlled trial. BMJ open respiratory research 2020;7(1):e000737.

42. Bittman B, Poornima I, Smith MA, Heidel RE. Gospel Music: A Catalyst for Retention, Engagement, and Positive Health Outcomes for African Americans in a Cardiovascular Prevention and Treatment Program. Advances in mind-body medicine 2020;34(1):8–16.

43. Innes KE, Selfe TK, Kandati S, Wen S, Huysmans Z. Effects of mantra meditation versus music listening on knee pain, function, and related outcomes in older adults with knee osteoarthritis: an exploratory Randomized Clinical Trial (RCT). Evidence-Based Complementary and Alternative Medicine 2018;2018.

44. Innes KE, Selfe TK, Khalsa DS, Kandati S. Effects of meditation versus music listening on perceived stress, mood, sleep, and quality of life in adults with early memory loss: A pilot randomized controlled trial. Journal of Alzheimer’s Disease 2016;52(4):1277–1298.

45. Lavretsky H, Epel E, Siddarth P, et al. A pilot study of yogic meditation for family dementia caregivers with depressive symptoms: effects on mental health, cognition, and telomerase activity. International journal of geriatric psychiatry 2013;28(1):57–65.

46. Mandel SE, Hanser SB, Secic M, Davis BA. Effects of music therapy on health-related outcomes in cardiac rehabilitation: a randomized controlled trial. Journal of Music Therapy 2007;44(3):176–197.

47. Warkentin L, Das D, Majumdar S, Johnson J, Padwal R. The effect of weight loss on health-related quality of life: systematic review and meta-analysis of randomized trials. Obesity Reviews 2014;15(3):169–182.

48. Hart PD, Buck DJ. The effect of resistance training on health-related quality of life in older adults: Systematic review and meta-analysis. Health promotion perspectives 2019;9(1):1.

49. Tanaka R, Ozawa J, Kito N, Moriyama H. Does exercise therapy improve the health-related quality of life of people with knee osteoarthritis? A systematic review and meta-analysis of randomized controlled trials. Journal of physical therapy science 2015;27(10):3309–3314.

50. Burger JP, de Brouwer B, IntHout J, Wahab PJ, Tummers M, Drenth JP. Systematic review with meta-analysis: Dietary adherence influences normalization of health-related quality of life in coeliac disease. Clinical nutrition 2017;36(2):399–406.

51. Regensteiner JG, Ware Jr JE, McCarthy WJ, et al. Effect of cilostazol on treadmill walking, community-based walking ability, and health-related quality of life in patients with intermittent claudication due to peripheral arterial disease: meta-analysis of six randomized controlled trials. Journal of the American Geriatrics Society 2002;50(12):1939–1946.

52. Arcoverde FVL, de Paula Andres M, Borrelli GM, de Almeida Barbosa P, Abrão MS, Kho RM. Surgery for endometriosis improves major domains of quality of life: a systematic review and meta-analysis. Journal of minimally invasive gynecology 2019;26(2):266–278.

53. Szmulewicz A, Wanis KN, Gripper A, et al. Mental health quality of life after bariatric surgery: A systematic review and meta-analysis of randomized clinical trials. Clinical obesity 2019;9(1):e12290.

54. Koelsch S. A coordinate-based meta-analysis of music-evoked emotions. Neuroimage 2020;223:117350.

55. Lemstra M, Bird Y, Nwankwo C, Rogers M, Moraros J. Weight loss intervention adherence and factors promoting adherence: a meta-analysis. Patient preference and adherence 2016;10:1547.

56. Schutzer KA, Graves BS. Barriers and motivations to exercise in older adults. Preventive medicine 2004;39(5):1056–1061.

57. Gold BP, Mas-Herrero E, Zeighami Y, Benovoy M, Dagher A, Zatorre RJ. Musical reward prediction errors engage the nucleus accumbens and motivate learning. Proceedings of the National Academy of Sciences 2019;116(8):3310–3315.

58. Fancourt D, Aughterson H, Finn S, Walker E, Steptoe A. How leisure activities affect health: a narrative review and multi-level theoretical framework of mechanisms of action. The Lancet Psychiatry 2021.

59. McCrary JM, Altenmüller E. Mechanisms of Music Impact: Autonomic Tone and the Physical Activity Roadmap to Advancing Understanding and Evidence-Based Policy. Frontiers in Psychology 2021:3790.

60. Cuijpers P, Weitz E, Cristea I, Twisk J. Pre-post effect sizes should be avoided in meta-analyses. Epidemiology and psychiatric sciences 2017;26(4):364–368.

61. Page MJ, McKenzie JE, Bossuyt PM, et al. The PRISMA 2020 statement: an updated guideline for reporting systematic reviews. Bmj 2021;372.

