## Supplementary Appendix for "The impact of music on health-related quality of life, as quantified by the SF-36: A systematic review and meta-analysis"

*Table of Contents*

***Table S1***. *GRADE Quality of Evidence ratings for included studies* 2

***Table S2***. *Articles excluded after full-text review, with reasons* 4

***Table S3***. *Reviews searched for additional records in this meta-analysis* 8

***Figure S1***. *Funnel plot detailing distribution of MCS pre-post changes* 10

***Figure S2***. *Funnel plot detailing distribution of PCS pre-post changes* 11

***Figure S3***. *Funnel plot detailing distribution of MCS changes in music+TAU vs. TAU interventions* 12

***Figure S4***. *Funnel plot detailing distribution of PCS changes in music+TAU vs. TAU interventions* 13

***Figure S5***. *Meta-analysis of MCS scores in music vs. meditation interventions* 14

***Figure S6***. *Meta-analysis of PCS scores in music vs. meditation interventions* 14

| Author | Year | Study Design | GRADE Criteria: *Intention to treat analysis* | *All outcome results reported* | *Blinding (assessors/data analysts)* | *Allocation concealment* | *% lost to follow-up*[*] | *Use of individual randomization* | *Controlled crossover effects* | **Overall quality rating** |
|---|---|---|---|---|---|---|---|---|---|---|
| Altena et al.[33] | 2009 | RCT | Yes | Yes | Yes | Yes | None | Yes | N/A | High |
| Ashok, Shanmugam & Soman[29] | 2019 | RCT | Yes | Yes | Unclear | Yes | Moderate (25%) | Yes | N/A | Moderate |
| Atiwannapat et al.[37] | 2016 | RCT | Yes | Yes | Yes | Unclear | Moderate (20%) | Yes | N/A | Moderate |
| Bittman et al. | 2020 | RCT | No (*analysis only on those who attended at least 12 sessions*) | Yes | No | Unclear | Unclear | Yes | N/A | Low |
| Bittman et al.[43] | 2020 | RCT | Yes | Yes | Yes | Unclear | Low (10%) | Yes | N/A | Moderate |
| Burrai et al.[30] | 2020 | Single group study | Yes | Yes | No | No | Low (9%) | N/A | N/A | Low |
| Corvo, Skingley & Clift[31] | 2015 | RCT | Yes | Yes | Unclear | Yes | Moderate (20%) | Yes | N/A | Moderate |
| Coulton et al.[39] | 2014 | Single group study | Yes | Yes | N/A | N/A | Moderate (19%) | N/A | N/A | Very low |
| Davidson et al.[23] | 2012 | Single group study | No (*those who missed 4+ rehearsals excluded*) | Yes | N/A | N/A | High (33%) | N/A | N/A | Very low |
| Gale et al.[40] | 2015 | RCT | Yes | Yes | Unclear | Unclear | None | Yes | N/A | Moderate |
| Groener et al.[28] | 2020 | Single group study | Yes | Yes | N/A | N/A | Moderate (26%) | N/A | N/A | Very low |
| Hagemann, Martin & Neme[24] | 2018 | RCT | Yes | Yes | Yes | Yes | None | Yes | N/A | High |
| Innes et al.[44] | 2016 | RCT | Yes | Yes | Yes | Yes | None | Yes | N/A | High |
| Innes et al.[45] | 2013 | RCT | Yes | No (*only MCS and selected subscales of SF-36 reported*) | Yes | Yes | Moderate (20%) | Yes | N/A | Moderate |
| Lavretsky et al.[46] | 2010 | RCT | Yes | Yes | No | Yes | Low (11%) | Yes | N/A | Moderate |
| Lee, Chan & Mok[27] | 2007 | RCT | Yes | Yes | Yes | Yes | None | Yes | N/A | High |
| Logtenberg et al.[34] | 2012 | RCT | Yes | Yes | Yes | Yes | Moderate (28%) | No (*randomized in blocks of 4*) | N/A | Moderate |
| Lord et al.[41] | 2007 | RCT | Yes | Yes | Unclear | Unclear | High (36%) | Yes | N/A | Low |

| | | | | | | | | | | |
|---|---|---|---|---|---|---|---|---|---|---|
| Mandel et al.[47] | 2018 | Single group study | Yes | Yes | Unclear | Unclear | None | Yes | Yes | Moderate |
| Mateu et al.[35] | 2015 | Single group study | Yes | Yes | N/A | N/A | None | N/A | N/A | Low |
| Mujdeci et al.[38] | 2020 | RCT | Yes | Yes | Yes | Yes | None | Yes | N/A | High |
| Philip et al.[42] | 2018 | RCT | No (*analysis only of those with >75% compliance*) | Yes | Unclear | Unclear | High | Yes | N/A | Low |
| Ribeiro[25] | 2020 | RCT | Yes | Yes | Unclear | Yes | High (34%) | No (*randomized in blocks of 6*) | N/A | Low |
| Wahlstöm et al.[36] | 2009 | RCT | Yes | Yes | Unclear | Unclear | None | Unclear | N/A | Low |
| Zanini et al.[26] | 2021 | RCT | No (*4 of 30 patients excluded for 'low compliance'*) | Yes | Yes | Yes | None | Yes | N/A | Moderate |

**Table S1.** Details of GRADE ratings of included studies.

| Author | Year | Title | Journal | Reason for Exclusion |
|---|---|---|---|---|
| Akmese & Oran | 2014 | Journal of Midwifery and Women's Health | Effects of Progressive Muscle Relaxation Exercises Accompanied by Music on Low Back Pain and Quality of Life During Pregnancy | Not a music-focused intervention |
| Ashok & Soman | 2018 | International Journal of Pharma and Bio Sciences | Efficacy of music therapy on hospital induced anxiety and health related quality of life in coronary artery bypass graft patients: Study protocol for a randomized controlled trial | Study protocol for Ashok, Shanmugam & Soman 2019 (included) |
| Bell, McIntyre & Hadley | 2016 | Psychomusicology: Music, Mind and Brain | Listening to classical music results in a positive correlation between spatial reasoning and mindfulness | No use of SF-36/12 |
| Bidabadi & Mehryar | 2015 | Journal of Affective Disorders | Music therapy as an adjunct to standard treatment for obsessive compulsive disorder and co-morbid anxiety and depression: A randomized clinical trial | No use of SF-36/12 |
| Bygren et al. | 2009 | Psychosomatic Medicine | Cultural participation and health: A randomized controlled trial among medical care staff | No compatible SF-36 data; SF-36 data only available as a composite of multiple interventions |
| Cao et al. | 2016 | International Journal of Clinical and Experimental Medicine | Music therapy improves pregnancy-induced hypertension treatment efficacy | No pre and post-test SF-36 data |
| Castelino et al. | 2013 | Australasian Psychiatry | The effect of group music therapy on anxiety, depression and quality of life in older adults with psychiatric disorders | No pre and post-test SF-36 data collection (data only at pre- and 4-weeks post-intervention completion) |
| Cohen et al. | 2006 | The Gerontologist | The Impact of Professionally Conducted Cultural Programs on the Physical Health, Mental Health, and Social Functioning of Older Adults | No use of SF-36/12 |
| Cooke et al. | 2010 | Aging & Mental Health | A randomized controlled trial exploring the effect of music on agitated behaviours and anxiety in older people with dementia | No use of SF-36/12 |
| Erkkila et al. | 2011 | British Journal of Psychiatry | Individual music therapy for depression: A randomised controlled trial | No compatible SF-36 data available |
| Erkkila et al. | 2021 | Frontiers in Psychology | Music Therapy for Depression Enhanced With Listening Homework and Slow Paced Breathing: A Randomised Controlled Trial | No compatible SF-36 data available |
| Franco et al. | 2014 | Psychology of Music | Affect-matching music improves cognitive performance in adults and young children for both positive and negative emotions | No use of SF-36/12 |
| Gold et al. | 2013 | Psychotherapy & Psychosomatics | Individual music therapy for mental health care clients with low therapy motivation: Multicentre randomised controlled trial | No use of SF-36/12 |
| Hattori et al. | 2011 | Geriatrics & Gerontology International | Controlled study on the cognitive and psychological effect of coloring and drawing in mild Alzheimer's disease patients | Not a music-focused intervention |
| Heiderscheit | 2006 | Thesis | The effects of the Bonny Method of Guided Imagery and Music on interpersonal problems, sense of coherence and salivary immunoglobulin a of adults in chemical dependency treatment | No use of SF-36/12 |
| Henneghan & Becker | 2019 | Archives of Physical Medicine and Rehabilitation | Improving Cognitive and Psychosocial Symptoms and Social Functioning in Breast Cancer Survivors | No compatible SF-36 data available in conference abstract (social subscale reported only) |
| Hofmann et al. | 2010 | 17th International Congress on Sound & Vibration | Additional effects of multisensory perception of music with a vibroacoustic mat to pure listening of music | No use of SF-36/12 (use of modified SF-12) |

| | | | | |
|---|---|---|---|---|
| Hseih et al. | 2019 | European Journal of Cancer Care | Effect of home-based music intervention versus ambient music on breast cancer survivors in the community: A feasibility study in Taiwan | No use of SF-36/12 |
| Innes et al. | 2018 | Journal of Alzheimer's Disease | Effects of meditation and music-listening on blood biomarkers of cellular aging and Alzheimer’s disease in adults with subjective cognitive decline: An exploratory randomized clinical trial | Another report of study described in Innes et al. 2016 (included) with fewer participants |
| Innes et al. | 2016 | Complementary Therapies in Medicine | A randomized controlled trial of two simple mind-body programs, Kirtan Kriya meditation and music listening, for adults with subjective cognitive decline: Feasibility and acceptability | Another report of study described in Innes et al. 2018 (included) |
| Jeon, Kim & Yoo | 2009 | Journal of Korean Academy of Nursing | Effects of music therapy and rhythmic exercise on quality of life, blood pressure and upper extremity muscle strength in institution-dwelling elderly women | No compatible SF-36 data available |
| Kim & Kang | 2021 | Geriatric Nursing | Effect of a group music intervention on cognitive function and mental health outcomes among nursing home residents: A randomized controlled pilot study | No use of SF-36/12 |
| Liddle, Parkinson & Sibbritt | 2012 | Australasian Journal on Ageing | Painting pictures and playing musical instruments: Change in participation and relationship to health in older women | Not an intervention study |
| Lin et al. | 2020 | Annals of Thoracic and Cardiovascular Surgery | Effect of Music Therapy on the Chronic Pain and Midterm Quality of Life of Patients after Mechanical Valve Replacement | No pre and post-test SF-36 data |
| Lin et al. | 2020 | Annals of Thoracic and Cardiovascular Surgery | Effect of Music Therapy on the Chronic Pain and Midterm Quality of Life of Patients after Mechanical Valve Replacement | No pre and post-test SF-36 data |
| Lord et al. | 2010 | American Journal of Respiratory and Critical Care Medicine | Effect of singing lessons in patients with COPD - A randomised controlled trial | Abstract version of Lord et al. 2012 (included) |
| Lord et al. | 2012 | American Journal of Respiratory and Critical Care Medicine | Effects of "singing for breathing" TM in patients with chronic obstructive pulmonary disease (COPD)-a randomized control trial | Abstract version of Lord et al. 2012 (included) |
| Lord et al. | 2011 | Journal of Aerosol Medicine and Pulmonary Drug Delivery | Singing for breathing effects of singing lessons in patients with COPD-a randomised control trial | Another report of study described in Lord et al. 2012 (included) |
| Lord et al. | 2010 | BMC Pulmonary Medicine | Singing teaching as a therapy for chronic respiratory disease--a randomised controlled trial and qualitative evaluation | Another report of study described in Lord et al. 2012 (included) |
| Low et al. | 2020 | Journal of Alternative and Complementary Medicine | Vocal music therapy for chronic pain: A mixed methods feasibility study | No use of SF-36/12 |
| Mandel et al. | 2007 | Journal of Music Therapy | Effects of music therapy on health-related outcomes in cardiac rehabilitation: A randomized controlled trial | No pre and post-test SF-36 data |
| Mandel, Davis & Secic | 2014 | Hospital Topics | Effects of music therapy on patient satisfaction and health-related quality of life of hospital inpatients | No pre and post-test SF-36 data |
| Mateu et al. | 2012 | Basic and Clinical Pharmacology and Toxicology | Jacobson's progressive muscle relaxation as adjunctive therapy in osteoarticular chronic pain | Abstract version of Mateu et al. 2018 (included) |
| Novotna et al. | 2017 | Multiple Sclerosis | Effect of music therapy on common symptoms of multiple sclerosis | No compatible SF-36 data available in conference abstract |
| Pearce et al. | 2016 | Journal of Community & Applied Social Psychology | Is group singing special? Health, well-being and social bonds in community-based adult education classes | No use of SF-36/12 (use of modified SF-36) |
| Poćwierz-Marciniak & Bidzan | 2017 | Health Psychology Report | The influence of music therapy on quality of life after a stroke | No compatible SF-36 data available |

| | | | | |
|---|---|---|---|---|
| Puhan et al. | 2006 | BMJ | Didgeridoo playing as alternative treatment for obstructive sleep apnea syndrome: randomised controlled trial | No pre and post-test SF-36 data |
| Raglio et al. | 2016 | International Journal of Neuroscience | Improvement of spontaneous language in stroke patients with chronic aphasia treated with music therapy: A randomized controlled trial | No compatible SF-36 data available |
| Reagon et al. | 2017 | European Journal of Cancer Care | Choir singing and health status in people affected by cancer | Observational, not intervention study |
| Ross, Hollen & Fitzgerald | 2006 | American Journal of Kidney Disease | Observational study of an Arts-in-Medicine Program in an outpatient hemodialysis unit | Not a music-focused intervention |
| Ross, Hollen & Fitzgerald | 2006 | American Journal of Kidney Disease | Observational study of an Arts-in-Medicine Program in an outpatient hemodialysis unit | Observational, not intervention study |
| Russ et al. | 2020 | Journal of Alternative and Complementary Medicine | Cortisol as an acute stress biomarker in young hematopoietic cell transplant patients/caregivers: Active music engagement protocol | No use of SF-36/12 |
| Shiranbidabadi & Mehryar | 2015 | Journal of Affective Disorders | Music therapy as an adjunct to standard treatment for obsessive compulsive disorder and co-morbid anxiety and depression: A randomized clinical trial | No use of SF-36/12 |
| Skingley et al. | 2011 | BMC Public Health | The effectiveness and cost-effectiveness of a participative community singing programme as a health promotion initiative for older people: protocol for a randomised controlled trial | Another report of study described in Coulton et al. 2015 (included) |
| Skingley et al. | 2014 | Arts & Health | Singing for breathing: Participants' perceptions of a group singing programme for people with COPD | Another report of study described in Coulton et al. 2015 (included) |
| Skingley et al. | 2014 | Arts & Health | Singing for breathing: Participants' perceptions of a group singing programme for people with COPD | No reporting of SF-36 results |
| Skingley, Marin & Clift | 2016 | Journal of Applied Gerontology | The contribution of community singing groups to the well-being of older people: Participant perspectives from the United Kingdom | Another report of study described in Coulton et al. 2015 (included) |
| Tai, Wang & Yang | 2015 | Neuropsychiatric Disease and Treatment | Effect of music intervention on the cognitive and depression status of senior apartment residents in Taiwan | No use of SF-36/12 |
| Unspecified (https://trialsearch.who.int/Trial2.aspx?TrialID=ACTRN12616001671459) | 2016 | ACTRN Clinical Trials Registry | Personalised relaxation practice to improve sleep quality in patients with chronic fatigue syndrome and depression: a Randomised Control Trial | No study results available |
| Unspecified (https://trialsearch.who.int/Trial2.aspx?TrialID=ISRCTN50156343) | 2012 | ISRCTN | Music and expressive arts therapy for women with a history of gynaecological cancer | No use of SF-36/12 |
| Unspecified (http://www.who.int/trialsearch/Trial2.aspx?TrialID=ACTRN12614000168651) | 2014 | ACTRN Clinical Trials Registry | Music Therapy for Older Adults | Not a music-focused intervention |
| Unspecified (http://www.who.int/trialsearch/Trial2.aspx?TrialID=DRKS00024549) | 2021 | DRKS (clinical trials registry) | Effects of Receptive Music Therapy with a Monochord in multiple sclerosis (MUTIMS) – a randomized controlled study | Study ongoing |

| | | | | |
|---|---|---|---|---|
| Unspecified (http://www.who.int/trialsearch/Trial2.aspx?TrialID=ISRCTN42943709) | 2019 | ISRCTN | Singing and COPD: a pilot randomised controlled trial | Study ongoing |
| Unspecified (https://clinicaltrials.gov/show/NCT00500526) | 2007 | Clinicaltrials.gov | Effects of Singing in Chronic Obstructive Pulmonary Disease | No use of SF-36, confirmed through author contact |
| Unspecified (https://clinicaltrials.gov/show/NCT03076801) | 2017 | Clinicaltrials.gov | Does Choral Singing Help Improve Stress in Patients With Ischemic Heart Disease? | No study results available |
| Unspecified (https://clinicaltrials.gov/show/NCT04034212) | 2019 | Clinicaltrials.gov | Singing for Health: improving Experiences of Lung Disease (SHIELD Trial) | Study ongoing |
| Unspecified (https://clinicaltrials.gov/show/NCT04446624) | 2020 | Clinicaltrials.gov | Oxidative Stress, Anxiety and Depression in Breast Cancer Patients: impact of Music Therapy | Not a music-focused intervention |
| Unspecified (https://clinicaltrials.gov/show/NCT04638244) | 2020 | Clinicaltrials.gov | Brief Online Music Intervention (BOMI) in Improving the Mental Well-being of Young People in the Community in Hong Kong | Study ongoing |
| Vara et al. | 2020 | International Urogynecology Journal | Music therapy in rehabilitation treatment for chronic pelvic pain | No compatible SF-36 data available in conference abstract |
| Wahlstöm et al. | 2018 | Circulation | Mediyoga improves health related quality of life and blood pressure among patients with paroxysmal atrial fibrillation-the MYPAF study | Abstract version of Wählstrom et al. 2020 (included) |
| Zanini et al. | 2010 | Journal of Hypertension | Music therapy contributing to the quality of life of hypertensive patients | Abstract version of Zanini et al. 2009 (included) |
| Zheng & Zhang | 2020 | Basic and Clinical Pharmacology and Toxicology | Effect of Music on Novel Coronavirus Pneumonia Patients' Rehabilitation Training after Recovery | No compatible SF-36 data available in conference abstract |

**Table S2.** Articles excluded after full-text review, with reasons.

| Author | Year | Journal | Title |
|---|---|---|---|
| Archer, Buxton & Sheffield | 2015 | Psycho-Oncology | The effect of creative psychological interventions on psychological outcomes for adult cancer patients: A systematic review of randomised controlled trials |
| Bradt & Dileo | 2014 | Cochrane Database of Systematic Reviews | Music interventions for mechanically ventilated patients |
| Bradt et al. | 2016 | Cochrane Database of Systematic Reviews | Music interventions for improving psychological and physical outcomes in cancer patients |
| Bradt, Dileo & Potvin | 2013 | Cochrane Database of Systematic Reviews | Music for stress and anxiety reduction in coronary heart disease patients |
| Campbell, Bodkin-Allen & Swain | 2021 | Journal of Health Psychology | Group singing improves both physical and psychological wellbeing in people with and without chronic health conditions: A narrative review |
| Geretsegger et al. | 2014 | Cochrane Database of Systematic Reviews | Music therapy for people with autism spectrum disorder |
| Geretsegger et al. | 2017 | Cochrane Database of Systematic Reviews | Music therapy for people with schizophrenia and schizophrenia-like disorders |
| Jespersen et al. | 2015 | Cochrane Database of Systematic Reviews | Music for insomnia in adults |
| McNamaral et al. | 2017 | Cochrane Database of Systematic Reviews | Singing for adults with chronic obstructive pulmonary disease (COPD) |
| Sereda et al. | 2018 | Cochrane Database of Systematic Reviews | Sound therapy (using amplification devices and/or sound generators) for tinnitus |
| Sinha et al. | 2011 | Cochrane Database of Systematic Reviews | Auditory integration training and other sound therapies for autism spectrum disorders (ASD) |
| van der Steen et al. | 2018 | Cochrane Database of Systematic Reviews | Music-based therapeutic interventions for people with dementia |
| Galaal et al. | 2011 | Cochrane Database of Systematic Reviews | Interventions for reducing anxiety in women undergoing colposcopy |
| Halsbeck et al. | 2019 | Cochrane Database of Systematic Reviews | Musical and vocal interventions to improve neurodevelopmental outcomes for preterm infants |
| Aalbers et al. | 2017 | Cochrane Database of Systematic Reviews | Music therapy for depression |
| Ghetti et al. | 2020 | Cochrane Database of Systematic Reviews | Music therapy for people with substance use disorders |
| Irons et al. | 2019 | Cochrane Database of Systematic Reviews | Singing for people with Parkinson's disease |
| Irons, Kenny & Chang | 2010 | Cochrane Database of Systematic Reviews | Singing for children and adults with bronchiectasis |
| Irons et al. | 2019 | Cochrane Database of Systematic Reviews | Singing as an adjunct therapy for children and adults with cystic fibrosis |
| Leckey | 2011 | Journal of Psychiatric and Mental Health Nursing | The therapeutic effectiveness of creative activities on mental well-being: A systematic review of the literature |
| Lee et a. | 2015 | Chest | Distractive Auditory Stimuli in the Form of Music in Individuals With COPD A Systematic Review |

| | | | |
|---|---|---|---|
| Lin et al. | 2019 | Journal of Clinical Medicine | Music interventions for anxiety in pregnant women: A systematic review and meta-analysis of randomized controlled trials |
| Magee et al. | 2017 | Cochrane Database of Systematic Reviews | Music interventions for acquired brain injury |
| Phillip, Lewis & Hopkinson | 2019 | Breathe | Music and dance in chronic lung disease |

**Table S3.** Review articles included at full-text article review stage. Citations of these reviews were searched for additional relevant articles.

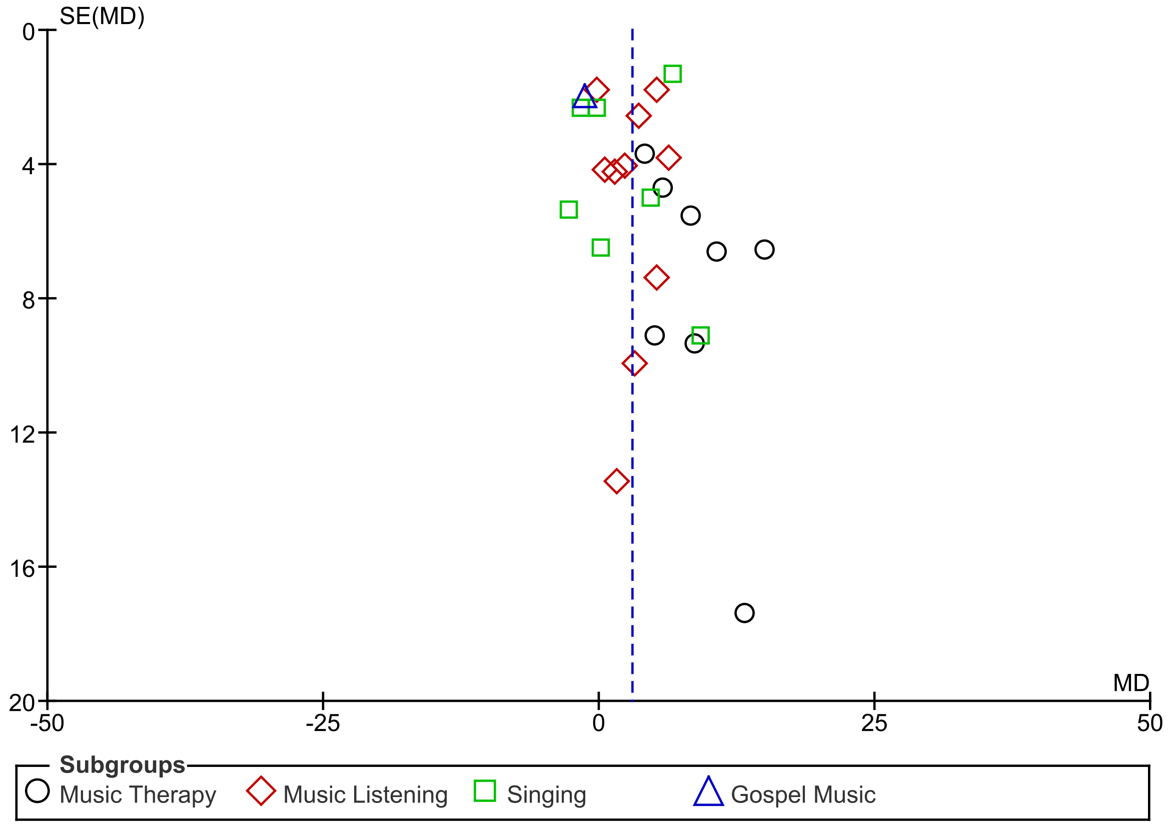


**Figure S1.** Funnel plot detailing the distribution of pre-post intervention changes in MCS score, stratified by music intervention type. SE(MD) = Standard error of the mean difference. MD = Mean difference.

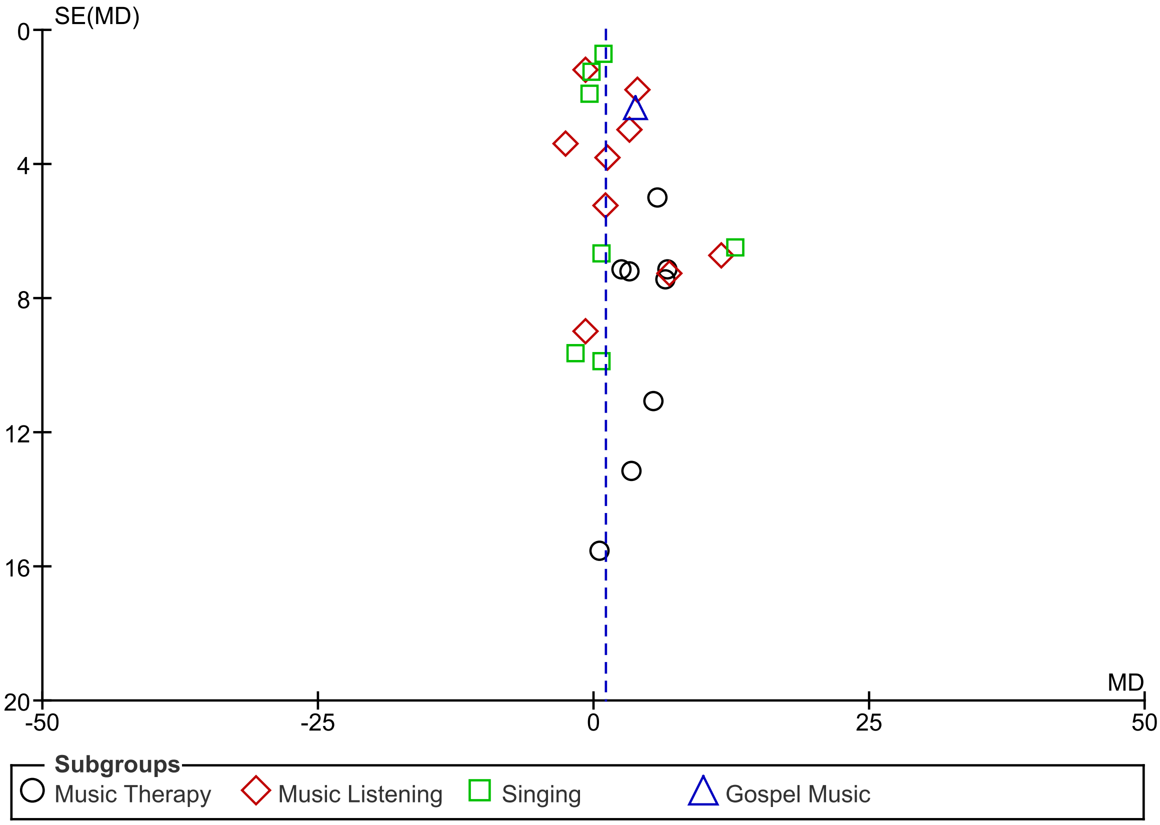


**Figure S2.** Funnel plot detailing the distribution of pre-post intervention changes in PCS score, stratified by music intervention type. SE(MD) = Standard error of the mean difference. MD = Mean difference.

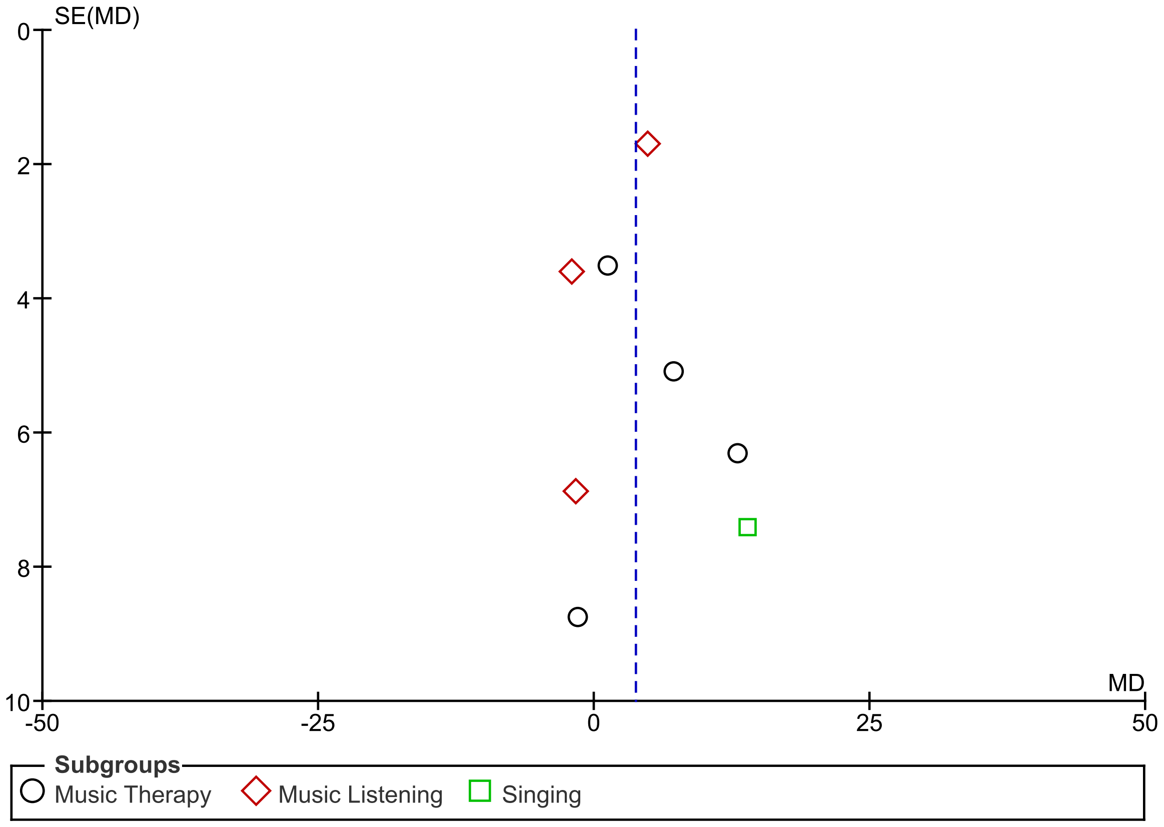


**Figure S3.** Funnel plot detailing the distribution of changes in MCS scores in Music+TAU vs. TAU alone interventions, stratified by music intervention type. SE(MD) = Standard error of the mean difference. MD = Mean difference.

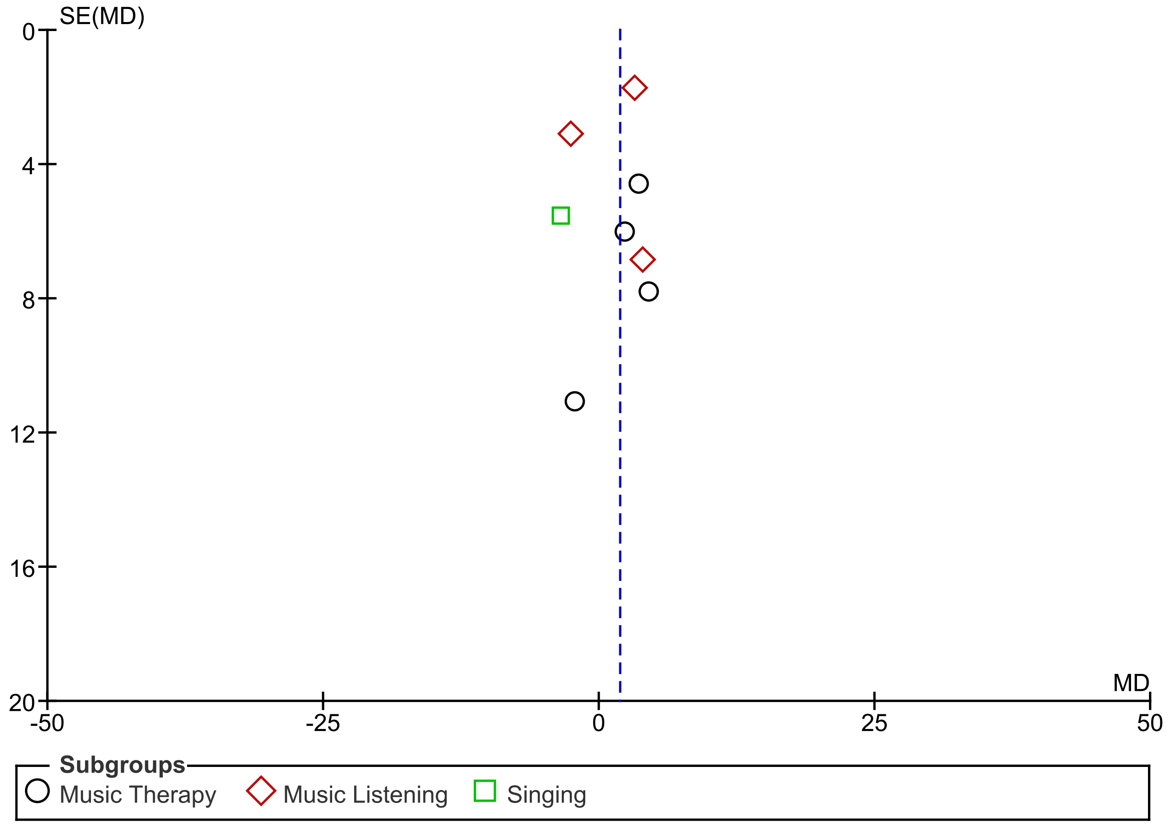


**Figure S4.** Funnel plot detailing the distribution of changes in PCS scores in Music+TAU vs. TAU alone interventions, stratified by music intervention type. SE(MD) = Standard error of the mean difference. MD = Mean difference.

| Study or Subgroup | Music Listening Mean | Music Listening SD | Music Listening Total | Meditation Mean | Meditation SD | Meditation Total | Weight | Mean Difference IV, Random, 95% CI |
|---|---|---|---|---|---|---|---|---|
| Innes et al. 2016 | 75.3 | 12 | 11 | 78.2 | 18.6 | 11 | 12.9% | -2.90 [-15.98, 10.18] |
| Innes et al. 2018 | 51.1 | 15.6 | 30 | 51.7 | 15.6 | 30 | 35.4% | -0.60 [-8.49, 7.29] |
| Lavretsky et al. 2013 | 43.6 | 10.6 | 16 | 47.2 | 9.7 | 23 | 51.7% | -3.60 [-10.13, 2.93] |
| **Total (95% CI)** | | | **57** | | | **64** | **100.0%** | **-2.45 [-7.15, 2.25]** |

Heterogeneity: $Tau^2 = 0.00$; $Chi^2 = 0.33$, df = 2 (P = 0.85); $I^2 = 0\%$
Test for overall effect: Z = 1.02 (P = 0.31)

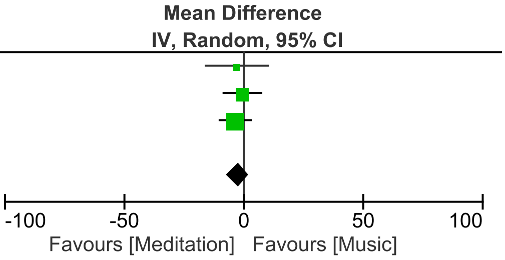


**Figure S5.** Meta-analysis of effects of music vs. meditation on SF-36 MCS scores. IV = ‘inverse variance’. ‘Total’ refers to the total number of participants included in analyses at pre- and post-intervention timepoints.

| Study or Subgroup | Music Listening Mean | Music Listening SD | Music Listening Total | Meditation Mean | Meditation SD | Meditation Total | Weight | Mean Difference IV, Random, 95% CI |
|---|---|---|---|---|---|---|---|---|
| Innes et al. 2016 | 59.5 | 12.5 | 11 | 66.4 | 17 | 11 | 34.9% | -6.90 [-19.37, 5.57] |
| Innes et al. 2018 | 46.5 | 16.6 | 30 | 45.9 | 19.4 | 30 | 65.1% | 0.60 [-8.54, 9.74] |
| **Total (95% CI)** | | | **41** | | | **41** | **100.0%** | **-2.02 [-9.39, 5.35]** |

Heterogeneity: $Tau^2 = 0.00$; $Chi^2 = 0.90$, df = 1 (P = 0.34); $I^2 = 0\%$
Test for overall effect: Z = 0.54 (P = 0.59)

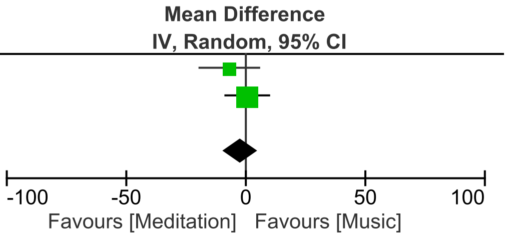


**Figure S6.** Meta-analysis of effects of music vs. meditation on SF-36 PCS scores. IV = ‘inverse variance’. ‘Total’ refers to the total number of participants included in analyses at pre- and post-intervention timepoints. NB: Lavretsky et al. 2013 reported MCS scores only.
